# Machine Learning-Based Pixel-Level Quantification of Intramuscular Connective Tissue using Ultrasound Texture Analysis

**DOI:** 10.1101/2024.08.21.24312346

**Authors:** Patricio A. Pincheira, Jong H. Kim, Paul W. Hodges

## Abstract

**Objective:** This study aimed to develop a machine learning method for characterizing muscle composition on ultrasound imaging, focusing on pixel-level quantification of connective tissue using texture analysis.

**Methods:** Ultrasound images of the multifidus muscle from 20 healthy young adults were included in the analysis. Texture features including Local Binary Patterns, Histograms of Oriented Gradients, Grey Level Co-occurrence Matrix, and Discrete Wavelet Transforms, were extracted from the images across multiple scales. Within a positive-unlabeled machine learning framework, two competing models, Bagging Support Vector Machine and Random Forests with Recursive Greedy Risk Minimization were trained for each texture and scale. The outputs of the texture-based pixel-level classification were compared to traditional echo intensity-based methods. Metrics such as the F-measure were employed to evaluate the models’ performance. Expert consensus was utilised to evaluate the accuracy of the classified images and identify the best-performing combination of model, texture, and scale.

**Results:** Expert evaluation identified the Bagging Support Vector Machine model trained with Local Binary Pattern histograms extracted at a scale of 9×9 pixel region of interest as the best combination for accurately classifying connective tissue-like pixels (F-measure= 0.88). The proposed method demonstrated high repeatability (intraclass correlation coefficient= 0.92) and robustness to echo intensity variations, outperforming traditional echo intensity-based methods.

**Conclusion:** This approach offers a valid method for pixel-level quantification of intramuscular connective tissue from ultrasound images. It overcomes the limitations of traditional analyses relying on echo intensity and demonstrates robustness against variations in echo intensity, representing an operator-independent advancement in ultrasound-based muscle composition analysis.

## Introduction

Muscle composition refers to the arrangement and proportion of muscle fibres, connective tissue, adipose, and other components within muscles. It is well known that muscle composition can be changed by a range of exposures and muscle pathologies. Amongst the many relevant examples, exercise and strength training change the quantity of contractile tissue within the muscle (1); different healing stages are associated with varying amounts of muscle fibrosis (i.e., connective tissue) (2); and muscular dystrophies are characterized by the loss of contractile tissue and infiltration of fatty tissue (3). Understanding the amount and distribution of these tissues within the muscle is crucial because muscle performance and function are largely determined by muscle structure. However, in vivo evaluation of muscle composition is not straightforward. Historically, computed tomography (CT) and magnetic resonance imaging (MRI) have been used (4). Yet, CT is limited by radiation exposure and limited soft tissue differentiation, while both methods are constrained by cost, availability, and long scan durations. Furthermore, most studies only quantified muscle mass and/or adipose tissue (4), without consideration of connective tissue. There is a need for more accessible, operationally simpler methods that can accurately quantify muscle composition, including connective tissues.

Ultrasound is extensively used to evaluate soft tissues such as muscle and tendon. Compared to other imaging techniques, ultrasound is safe, accessible, portable, and provides real-time imaging. Despite its potential to monitor musculoskeletal adaptations, sonography faces challenges that hamper its widespread use for muscle composition analysis. These challenges include high inter-operator variability and dependency of echo intensity on equipment settings (5–7). New image processing approaches that overcome the current limitations are needed to boost the application of muscle composition analysis using ultrasound imaging.

Texture analysis refers to a variety of mathematical and statistical methods that can be used to characterise the intensities and spatial distribution of pixels within an image (8). Higher-order texture features, which assess the relationships between 3 or more pixels within an image region of interest (RoI), offer surrogate metrics for evaluating muscle composition (5). This approach could provide better discriminatory value because is less influenced by muscle echo intensity variability. Although texture analyses are common for MRI (9), their use in ultrasound has been more recent and often focused on distinguishing different conditions (e.g., pathological vs. healthy) based on overall texture feature proportions (5). A detailed characterization of the quantity and distribution of different tissues within a muscle image requires a more detailed analysis at pixel level. This can be challenging because of the high-dimensional nature of texture features (5,8,9). In this regard, machine learning (ML) arises as a tool for automated analysis in ultrasonography (10). This approach has helped to accelerate image analysis by modelling complicated multidimensional data relationships to answer questions related to image classification and diagnosis (10). A ML model might help to classify the different textures (i.e., tissues) in an image, thus overcoming the current limitations in ultrasound image texture analysis.

The aim of this study was to refine a novel ML method to characterize muscle composition on ultrasound imaging, using pixel-level connective tissue quantification via texture analysis.

## Methods

### Overview of the proposed method

We developed a method leveraging texture feature descriptors to capture patterns between neighbouring pixels based on high spatial coherence, wherein a pixel is likely to exhibit similar properties to its neighbours (11,12). We explored diverse texture features across different dimensionalities, which were extracted and used in various ML models to predict the class (i.e., connective tissue) of each pixel within the muscle image (Figure 1). Based on classification quality, we then recommend the optimal parameter combination and model for the characterization of intramuscular connective tissue from ultrasound images.

**Figure 1.**
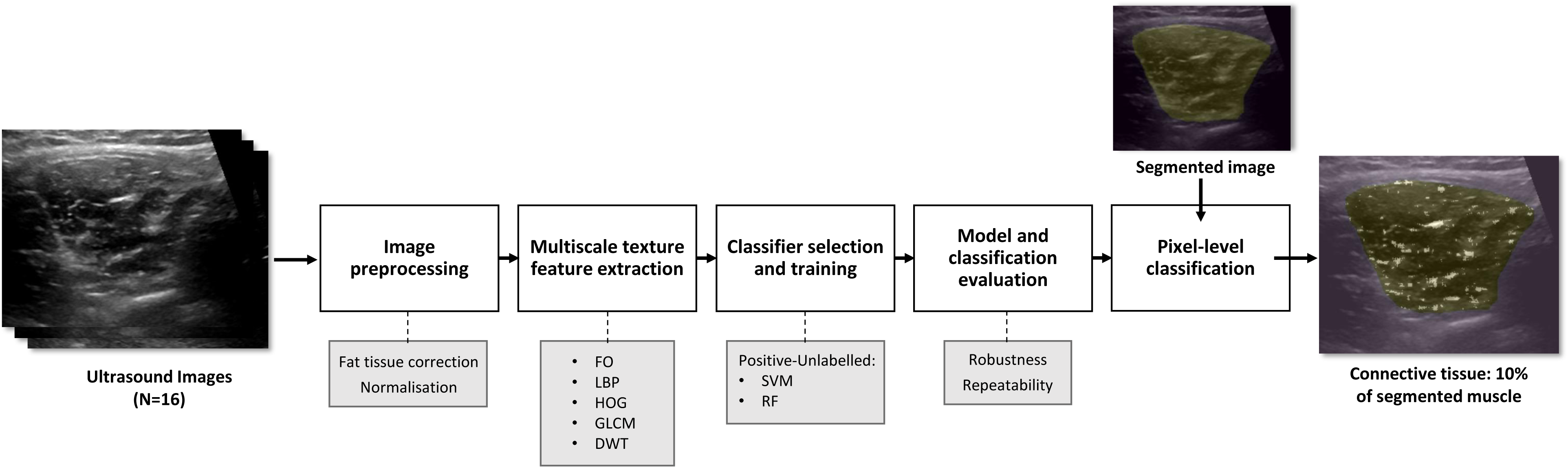
Overview of the proposed method. This method involves a series of steps starting with image preprocessing and multiscale texture feature extraction, finalising with an output image showing the distribution of pixels along with the proportion of classified pixels within the desired (segmented) muscle image. We tested five different textures – First Order Feature Vector, Local Binary Patterns, Histogram of Oriented Gradients, Gray Level Co-occurrence Matrix, and Discrete Wavelet Transform – (including multiple sub-options) and employed two different machine learning models for pixel classification – Support Vector Machine and Random Forest. The method’s classification robustness and repeatability were also assessed. Based on the classification quality, we recommend the best combination of parameters and models to be utilized with this method. See text for more details.

### Ultrasound images

For this study, we utilized a database of ultrasound recordings from a previous study (13) that had received approval from the University of Queensland’s Human Research Ethics Committee (ID: 2010000045). Twenty transverse B-mode ultrasound images (532 x 434 pixels, covering an area of 9 x 7.5 cm) from the right multifidus muscle at the fifth lumbar vertebrae were extracted. This muscle was selected because features of its composition have been shown to have clinical relevance in the prognosis and treatment of common spinal disorders (14). Ultrasound recordings were collected from twenty healthy young adults (10 male, age = 27 ± 7 years; weight = 67 ± 16 kg; height 170 ± 11 cm) using a LOGIQ 9 ultrasound system (GE, Milwaukee, WI), with a 10 MHz linear array transducer. All ultrasound settings were kept constant (frequency: 5 MHz, gain: 60), except for focus depth that was adjusted to the middle of the muscle for each participant. Recordings were made by evaluators who had been trained to image the multifidus muscle. Images were acquired with participants prone on a therapy table, with a pillow underneath their abdomen to decrease lumbar lordosis.

### Image preprocessing

To mitigate the impact of subcutaneous fat on image quality, we employed established equations to adjust pixel values according to the calculated thickness of adipose tissue within the image (15). Subsequently, grayscale images were normalized to enhance invariance to changes in focus depth and sonographers. This normalization involved linear scaling of pixel values between the local minimum and maximum of the image (16).

### Texture Features extraction

For texture feature extraction, the multifidus muscle was segmented by a single evaluator (J.H.K). This evaluator received training from the senior investigator, who has more than 20 years of experience in spine imaging analysis. All segmentations were performed manually using custom-made scripts written in Python (v3.9.12), using the Napari platform (17).

From each segmented image, we extracted four different higher-order (5) texture features: Local Binary Patterns (LBP), Histograms of Oriented Gradients (HOG), Grey Level Co-occurrence Matrix (GLCM), and Discrete Wavelet Transforms (DWT). These texture features differ in their underlying principles, the specific image characteristics they capture, and the types of spatial patterns they are designed to detect. Particularly, these textures have proven effective in estimating muscle composition from ultrasound images (5). A detailed description of each texture can be found elsewhere (5,18,19). We also calculated a custom first-order texture feature vector for each RoI, including echo intensity mean, standard deviation, skewness, kurtosis, median, entropy, and range. This descriptor, based solely on echo intensity values, allows direct comparison with texture features and has proven effective in pixel-level classification of carotid ultrasound images (11). Texture feature calculations were done using custom-made scripts written in Python, using the Scikit-image (18) and PyWavelets (19) libraries.

For multiscale texture feature extraction, a point outside the segmentation boundary was manually defined, assuming surrounding pixels belong to the thoracolumbar fascia which is composed of connective tissue (20). Seven pixel neighbourhoods (n×n, where n is an odd number between 3 and 15 pixels) were defined around this point. For each neighbourhood, corresponding RoIs of the same size of the neighbourhood were extracted for each pixel in the neighbourhood. The same process was repeated for every pixel within the boundaries of the segmented muscle.

### Classifier selection and training

Assuming muscle connective tissue shares a texture with thoracolumbar fascia, we aimed to identify each pixel within the segmented muscle with a connective tissue-like texture. Treating each pixel as a sample unit allowed us to extract sufficient data for ML model training. With only one known class (connective tissue-like texture) and uncertain identities for the remaining pixels (fat and contractile muscle tissue, both hyperechoic, are difficult to distinguish on ultrasound), we approached this as a binary classification problem with positive-unlabelled data.

Given that positive-unlabelled classification problems are uncommon in ML (21), we competed two models: Bagging Support Vector Machine (PU-SVM)(22) and Random Forests with Recursive Greedy Risk Minimization (PU-RF)(23). The PU-SVM iteratively trains binary classifiers on known positives and random unlabelled subsamples, averaging their predictions. The PU-RF recursively splits the dataset to minimize risk. These models, optimized for positive-unlabelled learning, were chosen for their simplicity, effectiveness, and proven success in prior binary classification tasks (24,25).

For model training, we separated the ultrasound images into a training dataset (16 images) and a test dataset (4 images). Positive data were texture features from pixels in the thoracolumbar fascia, while unlabelled data were pixels from the segmented muscle. Due to data imbalance, the unlabelled dataset was downsampled to ten times the size of the labelled dataset, with the process applied individually for each RoI scale. The training dataset size for each RoI size followed the formula: positive dataset = RoI size × RoI size × 16; unlabelled dataset = positive dataset × 10. That is, for instance, for a RoI size of 3, a positive dataset of 144 pixels and an unlabelled dataset of 1440 pixels were extracted. For each texture and RoI, the PU-SVM classifier parameters “C” and “γ” were tuned using a grid search, with performance evaluated by the area under the corrected precision-recall curve (26). Hyperparameter tuning was omitted for the PU-RF, as it is effective with default parameters (23).

ML models were trained for each combination of texture feature and RoI size. A detailed description of the feature descriptors employed during training is provided in Table 1. For textures with high dimensionality, Principal Component Analysis was employed for dimensionality reduction, retaining 90% of variance with 2 components, which were then used for model training. In total, 84 ML models were trained, encompassing 6 texture features, 7 RoI sizes, and 2 ML model types.

**Table 1.**
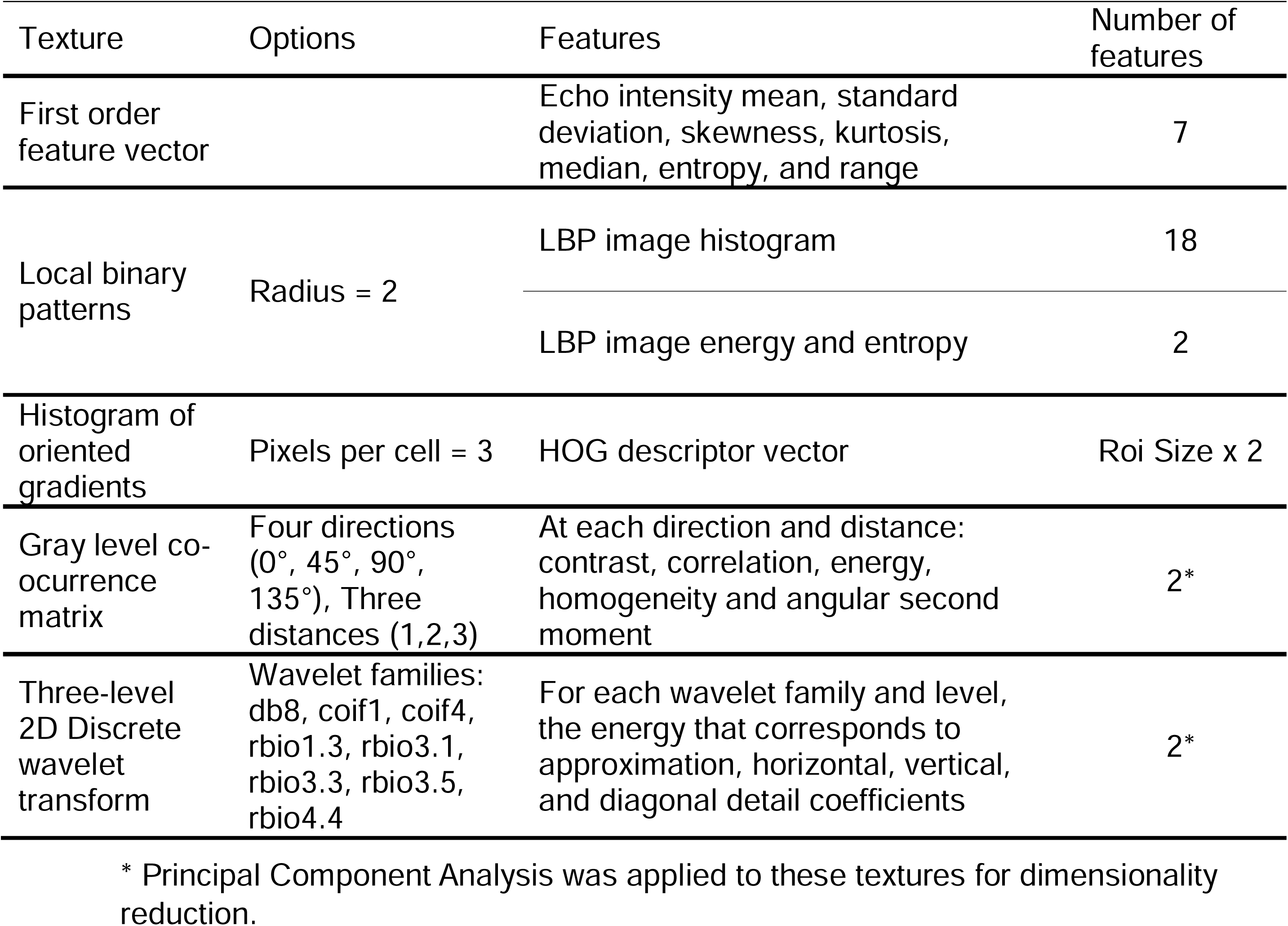
Texture feature descriptors used during machine learning model training.

| Texture | Options | Features | Number of features |
| --- | --- | --- | --- |
| First order feature vector |  | Echo intensity mean, standard deviation, skewness, kurtosis, median, entropy, and range | 7 |
| Local binary patterns | Radius = 2 | LBP image histogram | 18 |
|  |  | LBP image energy and entropy | 2 |
| Histogram of oriented gradients | Pixels per cell = 3 | HOG descriptor vector | Roi Size x 2 |
| Gray level co-occurrence matrix | Four directions (0°, 45°, 90°, 135°), Three distances (1,2,3) | At each direction and distance: contrast, correlation, energy, homogeneity and angular second moment | 2* |
| Three-level 2D Discrete wavelet transform | Wavelet families: db8, coif1, coif4, rbio1.3, rbio3.1, rbio3.3, rbio3.5, rbio4.4 | For each wavelet family and level, the energy that corresponds to approximation, horizontal, vertical, and diagonal detail coefficients | 2* |

\* Principal Component Analysis was applied to these textures for dimensionality reduction.

### Model and classification evaluation

Expert evaluation of classifier performance involved visual inspection of the output image that showcased the distribution of classified pixels. Four experts with more than 10 years of experience in musculoskeletal ultrasound imaging were presented with a set of classified images and were asked to identify the ML model and texture that produced images that most accurately represented the distribution of connective tissue in the ultrasound image. Specific attention was placed on identification of the method that performed well for both small and large areas of connective tissue with limited false positives (area identified as representing connective tissue that is unlikely to be connective tissue). Experts evaluated the images independently and a threshold for agreement was set at 3 of 4 assessors selecting the same combination of parameters.

To evaluate the performance of our models to select tissue that matches the target tissue (connective tissue from the thoracolumbar fascia), we calculated the F measure, precision, recall and Area under the Precision-recall curve on the pixels belonging to the test dataset. These metrics are considered appropriate for positive-unlabelled learning, given the class imbalance often present in such datasets (27). Because only positive and unlabelled examples were available, the standard calculation of these metrics is prohibited. Thus, we derived corrected (bounded) versions of these metrics (26), by using the distribution of decision values of known positives as a proxy for the distribution of decision values of all positives, assuming that the known positives are a representative (random) subset of all positives. Further, we determined empirical risk using zero-one loss as an indicator of model accuracy (23). This approach provides a reliable way to assess model performance with positive-unlabelled data (27). Each metric was calculated using a 3-fold cross validation process.

To assess the robustness of our texture-based classification to echo intensity variations, we generated synthetic images for each image in the test group. These synthetic images comprised versions with augmented and decreased echo intensity that were created by adjusting brightness/contrast by 20%. This approach simulated common B-mode ultrasound artifacts (e.g., shadowing and enhancement) on echo intensity (28). The classifier’s performance was subsequently compared to that of blob analysis (29, 30), identified as the most appropriate echo intensity-based muscle composition analysis. In brief, blob analysis involves examining groups of spatially connected pixels with similar intensities known as "blobs." In this process, a binary image of the segmented muscle is generated using a threshold based on mean pixel intensity from a reference image. Details of this calculation are provided elsewhere (5,29,30).

The outcome of the platform is an image that displays the distribution of classified pixels, and the calculation of the proportion of classified pixels in relation to the total number of pixels within the segmented image. To assess the repeatability of the quantification of connective tissue proportion, we classified four additional images. These images belonged to repeated ultrasound recordings of subjects from the test dataset. The repeated recordings were performed with the methods described above and were conducted with a three-week interval.

## Results

The results of this study indicate that the classification performance of each model depends on both the texture features and RoI size. Consensus among experts identified that the PU-SVM exhibited superior performance, particularly in detecting small clusters of pixels that were overlooked by the PU-RF. The performance of the PU-SVM model for the various textures and RoI sizes is illustrated in a classified image from a single representative participant of the test group (Figure 2). The performance of the PU-RF model for different textures and RoI sizes is presented in the Supplementary Material A.

**Figure 2:**
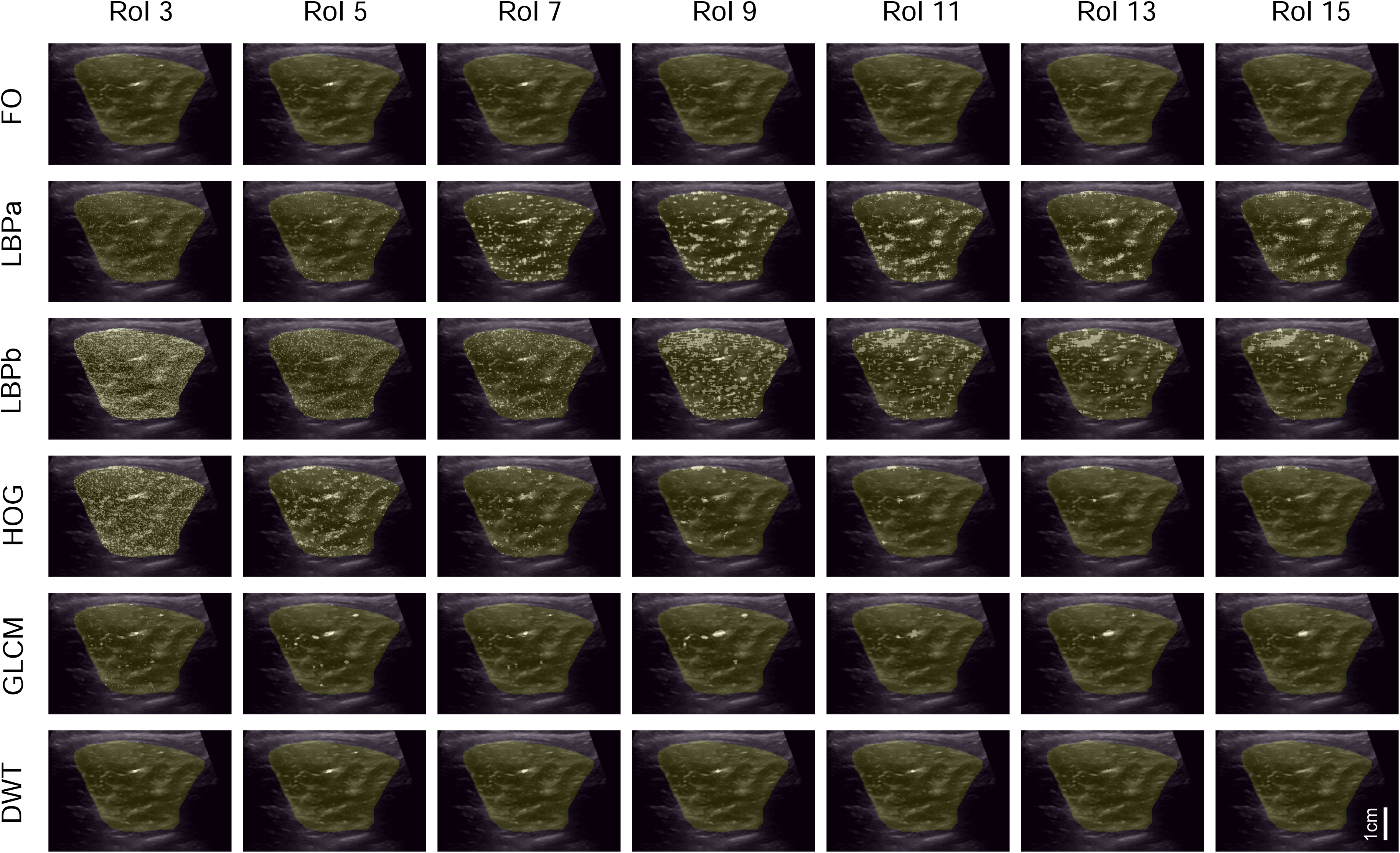
Classification outcomes for the positive-unlabelled support vector machine model. Different textures are presented in rows, region of interest (RoI) sizes in columns. The outcomes are illustrated in a classified image from a single representative participant. Segmented multifidus muscle is highlighted and white pixels within the segmented muscle represent pixels with texture classified as connective tissue. FO: First Order Feature Vector; LBPa: Local Binary Patterns image histogram; LBPb: Local Binary Patterns image energy and entropy; HOG: Histogram of Oriented Gradients; GLCM: Gray Level Co-occurrence Matrix; DWT: Discrete Wavelet Transform

Regarding texture, all four experts concurred that the texture with the best performance for selection of areas of connective tissue was the LBP histogram. Concerning RoI size, most experts favoured RoI 9 (three out of four). This combination of texture and RoI estimated that for the test dataset, the connective tissue texture within the muscle was 12%. Classified images for the test group based on the selected model, texture, and RoI size are shown in Figure 3. Performance metrics for the selected model, texture and RoI size are presented in Table 2.

**Figure 3:**
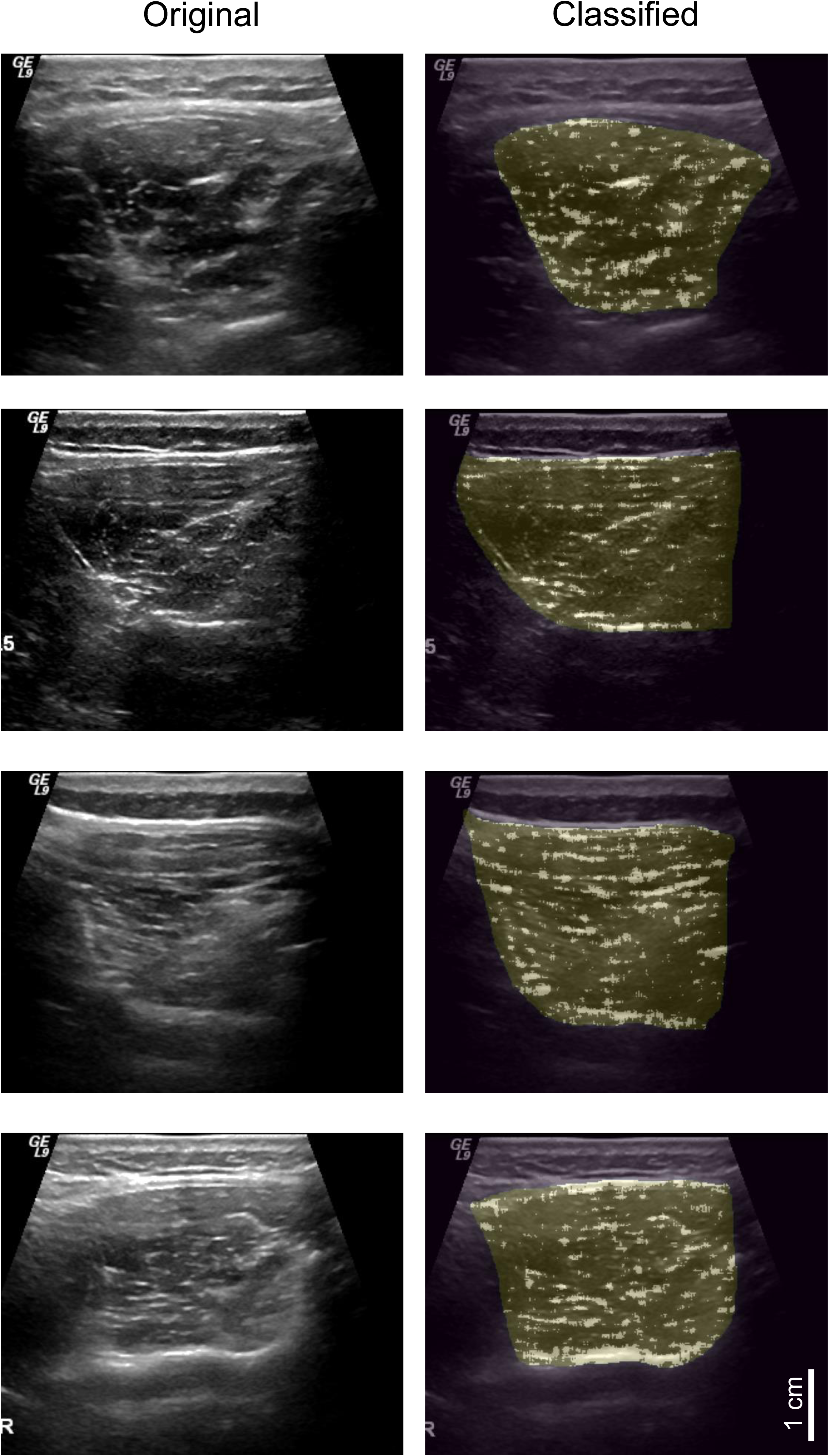
Classified images based on the preferred model (positive-unlabelled support vector machine, trained with Local Binary Patterns image histograms, extracted from 9×9 pixel region of interest). Original and classified images are shown in different columns. Each row represents a different participant from the test group. Segmented multifidus muscle is highlighted and white pixels within the segmented muscle represent pixels with texture classified as connective tissue.

**Table 2.** Model performance when selecting textures matching the connective tissue in the thoracolumbar fascia.

| Model | Texture<br>Parameters | F-measure | Precision | Recall | Area<br>under the<br>Precision-recall<br>curve | Empirical<br>risk using<br>zero-one<br>loss |
| --- | --- | --- | --- | --- | --- | --- |
| PU-SVM | LBP Image<br>histogram,<br>RoI 9 | 0.88 | 0.97 | 0.8 | 0.94 | 0.91 |

PU-SVM: Bagging Support Vector Machine; LBP: Local Binary Patterns; RoI: Region of interest. All metrics have been adjusted based on confidence intervals to account for the Positive-Unlabeled training (26).

In assessing the robustness of the chosen model, the analysis of synthetic images indicates that the proposed texture feature classifier is less susceptible to changes in echo intensity compared to blob metrics (Figure 4). The difference in the percentage of classified pixels with altered echo intensity was minimal with the PU-SVM classifier (1.6% and 1.9% for increased and decreased echo intensity, respectively). In contrast, this difference was notably higher for the blob analysis (41.6% and 4.7% for increased and decreased echo intensity, respectively). Furthermore, our method demonstrates excellent repeatability (intraclass correlation coefficient: 0.92, two-way mixed effects, absolute agreement, single rater/measurement (31)); Measurement 1: 11.2 ± 4.1%; Measurement 2: 11.9 ± 3.8%.

**Figure 4:**
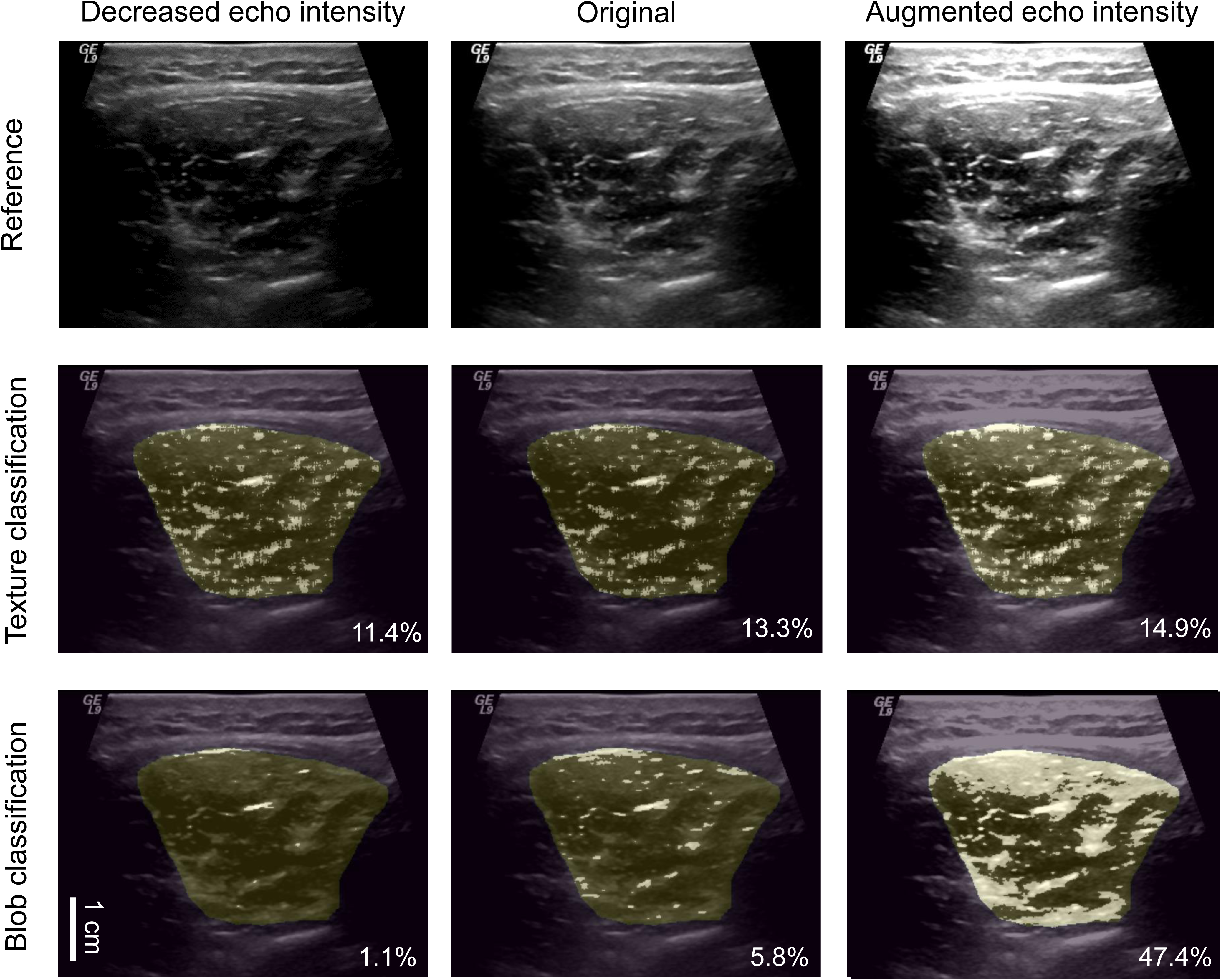
Classified synthetic images with augmented and decreased echo intensity. Original images and images with augmented and decreased echo intensity (20%) are shown in different columns. Reference images without classification and classified images using the preferred texture (positive-unlabelled support vector machine, trained with Local Binary Patterns image histograms, extracted from 9×9 pixel region of interest) and blob method are shown in rows. Segmented multifidus muscle is highlighted and white pixels within the segmented muscle represent pixels with texture classified as connective tissue. Numbers within the classified images represent the percentage of classified pixels in relation to the total number of pixels within the segmented muscle.

## Discussion

This study aimed to propose a ML-based method for pixel-level connective tissue quantification through texture analysis of ultrasound images. The approach, employing a PU-SVM model trained with LBP image histograms extracted with a RoI of size 9, successfully identified pixels that were considered to represent connective tissue in segmented multifidus ultrasound images. The resulting classification enables a comprehensive analysis of connective tissue distribution within the muscle, offering insights into its proportion relative to the entire muscle area. The method also demonstrates resilience against changes in echo intensity, representing a significant advancement that overcomes traditional limitations associated with quantification of muscle composition in ultrasound.

### Overall method considerations

To our knowledge, this is the first study to propose a method for the pixel-level quantification of connective tissue in ultrasound images. Our approach presents several advantages over previous methods that have predominantly relied on echo intensity as a practical surrogate measure for overall skeletal muscle composition (5). For instance, our method demonstrates efficacy with minimal susceptibility to changes in echo intensity. This robustness is attributed to the relative independence of texture features from echo intensity, allowing for its application irrespective of ultrasound settings/configuration or operator variability. Moreover, the efficiency of our classification is enhanced by analysis of the sample at the pixel level. This characteristic facilitates the data acquisition phase, requiring relatively few ultrasound images from different participants, as tens or hundreds of pixels can be extracted from a single image.

It is important to note that the model, texture, and RoI size exhibiting optimal performance here are likely to be specific to connective tissue. Given the unique characteristics of texture features within the ultrasound images, classification of a new tissue might require a different texture and RoI size. Future studies employing a similar workflow to that presented here may explore the classification of other type of tissues.

### Model selection

According to the opinion of the experts for this study, the PU-SVM demonstrated superior overall performance in the classification task. A key attribute of this model was the identification of small clusters of pixels overlooked by the PU-RF. This advantage may be related to the sensitivity of SVMs to detect local patterns in the data when utilizing a radial basis function (32). Because the ultrasound images contain small clusters with non-linear relationships or patterns, the SVM’s decision boundaries appear better equipped to capture them effectively. RFs construct multiple decision trees and aggregate their predictions through averaging or voting (23). Although this ensemble approach is potent and robust, it may encounter limitations in capturing very small, isolated clusters—especially if these clusters are inconsistently represented across the numerous “trees” in the “forest”. Nevertheless, we do not entirely dismiss the utility of the PU-RF model (or other models) in different scenarios (e.g. in situations where computational efficiency is crucial for optimization of the classification task).

### Texture selection

LBP histograms was found to be the best feature to represent the connective tissue texture. LBP is known for its robustness to changes in brightness and less sensitivity to noise compared to other texture descriptors (33). Given that ultrasound images often suffer from noise and speckle due to the imaging process, LBP may provide better discriminative features for the classification task. Further, its capacity to capture local patterns seems to be beneficial in describing texture variations within small regions of ultrasound images, potentially enhancing its effectiveness in highlighting specific structures or patterns relevant for classification. The superiority of the LBP histogram over LBP energy entropy can be attributed to the former’s capacity to capture the distribution of patterns in various spatial arrangements. In contrast, energy and entropy focus on quantification of degrees of diversity, randomness, and overall uniformity, qualities that may have diminished in our images because of the pre-processing step.

### Region of interest selection

We determined that RoI 9 yielded the most effective extraction of connective tissue-like features for the classification task. Other RoIs between 7-11 also demonstrated reasonable classification, indicating that this range of RoIs is suitable for representing the pixel’s spatial coherence (11,12) of connective tissue in ultrasound images. It is conceivable that the selection of RoIs might vary concerning specific details of the extracted texture. For example, LBP calculation depends on the radius parameter, encoding the relationship between the pixel of interest and its neighbours (33). We conducted preliminary analyses for these parameters across all textures, and pre-selected fixed parameters that better resembled what was expected for connective tissue. The impact of changing texture parameters on RoI selection and classification outcomes remains unknown.

### Limitations

The main limitation of the current method is that the biological significance of the pixels classified as having a connective tissue-like texture depended on expert opinion and requires further biological validation. Additional research is needed assess the validity of the classification in comparison to histologic or reference measures of intramuscular connective tissue. General agreement between the amount of connective tissue quantified here and findings from histological samples in other studies (34), supports the validity of our estimation.

Although ML performance metrics were reported for our model in the context of Positive-unlabelled data, it is important to note that these metrics may still be influenced by class imbalance, noisy labels from the unlabelled dataset, and feature importance. Thus, it is plausible that a model with superior classification outcomes captures essential information not adequately reflected in the chosen evaluation metrics (Supplementary Material B). Thus, contrary to sole reliance on these metrics, our model selection process followed an approach similar to that proposed by Pazinato et al. (11) for pixel-level classification of carotid images. In that method, an expert verified that the selected pixels belonged to the same tissue, making expert criteria the determinant of the ground truth. Although this method may be considered subjective, it produces results similar to the proportions provided by histological verification (11). In a scenario involving positive-unlabelled data with class imbalance and lacking ground truth for model evaluation, we prioritized the interpretability of the model’s decisions to choose the best model.

We used DICOM images directly exported from the ultrasound device. The impact of various image compression protocols (e.g., MP4) on the performance of the classification remains unknown. Additionally, the impact of changing probe frequency is unknown, but likely significant, as probe frequency affects image resolution and thus spatial coherence.

## Conclusion

This paper proposes a promising ML-based method for accurate pixel-level quantification and characterization of intramuscular connective tissue from ultrasound images. By employing a PU-SVM model trained on LBP image histograms extracted from 9×9 pixel ROIs, the approach effectively identifies connective tissue-like textures within the multifidus muscle.

This method overcomes limitations of traditional analyses that rely solely on echo intensity for muscle composition estimates. It is precise for delineation of small clusters of pixels classified as connective tissue. Further, its robustness to variations in echo intensity represents an operator-independent advancement towards mitigation of the impact of ultrasound artifacts on tissue characterization. With proper histological verification of the biological significance of classified pixel textures, this technique could enable investigations seeking to elucidate the relationship between muscle composition and function in health and disease.

## Acknowledgements

The authors would like to thank Jonathan Wilton for his assistance in implementing the positive-unlabelled random forest model.

## Conflict of Interest Statement

None to declare.

## Data Availability Statement

Data available upon reasonable request.

## Notes

### Competing Interest Statement

The authors have declared no competing interest.

### Funding Statement

This study did not receive any funding.

### Author Declarations

For this study we utilized a database of ultrasound recordings from a previous study that had received approval from the University of Queensland Human Research Ethics Committee (ID: 2010000045).

