## Supplementary figures and images for "Machine Learning-Based Pixel-Level Quantification of Intramuscular Connective Tissue using Ultrasound Texture Analysis"

### Supplementary Material A

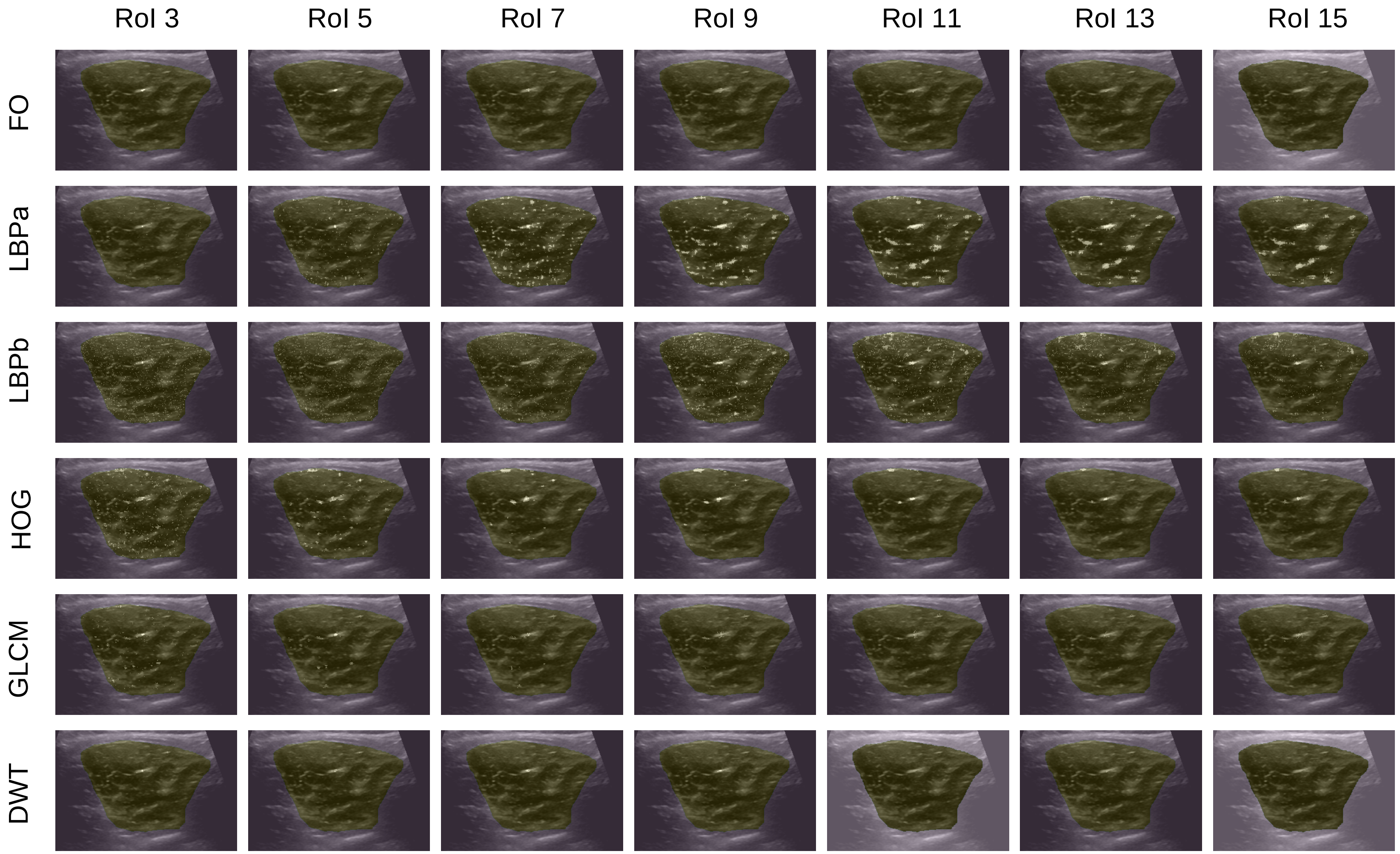
